# Deep Learning Spatial Profiling of CD103⁺CD8⁺ T Cells and Survival in Rectal Cancer After Neoadjuvant Chemoradiotherapy

**DOI:** 10.64898/2026.05.26.26353629

**Authors:** Tomoki Abe, Kimihiro Yamashita, Toru Nagasaka, Mitsugu Fujita, Yasuhiro Ueda, Soichiro Miyake, Ryota Ito, Yukari Adachi, Masayuki Ando, Takao Tsuneki, Yuki Okazoe, Ryunosuke Konaka, Toru Takahashi, Hiroki Kagiyama, Takaaki Tachibana, Masaki Imai, Takeshi Yoshida, Masafumi Saito, Junko Mukohyama, Kazuki Kanayama, Yu-Ichiro Koma, Yasunori Otowa, Hiroshi Hasegawa, Taro Ikeda, Yasufumi Koterazawa, Tomoaki Aoki, Hitoshi Harada, Naoki Urakawa, Hironobu Goto, Shingo Kanaji, Hiroaki Yanagimoto, Takeru Matsuda, Shiki Takamura, Tomoya Yamashita, Ryohei Sasaki, Takumi Fukumoto, Yoshihiro Kakeji

**Affiliations:** Division of Gastrointestinal Surgery, Department of Surgery, Graduate School of Medicine, Kobe University, Kobe, Japan; Division of Biophysics, Department of Health Sciences, Kobe University Graduate School of Medicine, Kobe, Japan; Association of Medical Artificial Intelligence Curation (AMAIC), Nagoya, Japan; Center for Medical Education and Clinical Training, Faculty of Medicine, Kindai University, Higashiosaka, Japan; Division of Hepato-Biliary and Pancreatic Surgery, Department of Surgery, Graduate School of Medicine, Kobe University, Kobe, Japan; Kobe University Graduate School of Science, Technology and Innovation, Kobe, Japan; Department of Immunology and Microbiology, National Defense Medical College, Tokorozawa, Japan; Department of Surgery, The Institute of Medical Science, The University of Tokyo, Minato-ku, Japan; Laboratory for Immunological Memory, RIKEN Center for Integrative Medical Science, Yokohama, Japan; Division of Radiation Oncology, Graduate School of Medicine, Kobe University, Kobe, Japan

**Keywords:** rectal cancer, neoadjuvant chemoradiotherapy, tumor-infiltrating lymphocytes, CD103⁺CD8⁺ T cells, tissue-resident memory-like T cells, deep learning, image cytometry

## Abstract

**Background:** CD8⁺ tumor-infiltrating lymphocytes (TILs) are established prognostic markers in colorectal cancer, yet the clinical significance of CD103⁺CD8⁺ tissue-resident memory–like (T_RM_-like) T cells in locally advanced rectal cancer (LARC) after neoadjuvant chemoradiotherapy (NACRT) remains unknown.

**Methods:** We quantified CD8⁺ and CD103⁺CD8⁺ T-cell densities in stromal and intratumoral compartments of post-NACRT resection specimens from 40 LARC patients using Cu-Cyto, a deep learning–based imaging cytometry platform. Associations with survival, pathological response, and adjuvant chemotherapy (AC) were examined. Treatment-induced T-cell dynamics were assessed in paired pretreatment biopsies and post-NACRT resections (*n* = 9).

**Results:** High stromal CD103⁺CD8⁺ density independently predicted better 5-year RFS (67.4% vs. 12.1%, *p* < 0.001) and OS (80.0% vs. 26.6%, *p* = 0.016); intratumoral density showed no prognostic significance. Pathological response correlated with stromal CD8⁺ but not CD103⁺CD8⁺ density. Paired analysis revealed a selective non-expansion of the CD103⁺ subset: stromal CD8⁺ T cells increased significantly after NACRT while CD103⁺CD8⁺ density remained unchanged. AC may preferentially benefit patients with low stromal CD103⁺CD8⁺ density.

**Conclusions:** Stromal CD103⁺CD8⁺ T-cell density is a robust independent prognostic biomarker in rectal cancer after NACRT that appears to reflect pre-existing rather than treatment-induced immunity. Given its stability across NACRT, pretreatment biopsy assessment may provide equivalent prognostic information, with potential implications for patient stratification before treatment initiation.

## Introduction

Locally advanced rectal cancer (LARC) is commonly treated with neoadjuvant chemoradiotherapy (NACRT) followed by radical surgery [1]. This multimodal strategy improves local control and facilitates sphincter preservation, but long-term outcomes remain heterogeneous, and a subset of patients still experience recurrence despite curative resection [1,2]. Reliable biomarkers that can stratify prognosis and guide postoperative treatment decisions are therefore urgently needed. The recent emergence of immune checkpoint blockade (ICB) combined with NACRT has brought renewed attention to the pre-existing tumor immune microenvironment as a potential determinant of both prognosis and treatment responsiveness.

Tumor-infiltrating lymphocytes (TILs), particularly CD8⁺ T cells, are key mediators of antitumor immunity. In rectal cancer, higher CD8⁺ T-cell densities have been associated with improved prognosis and treatment response after NACRT [3–6], although the strength and consistency of these associations vary. These findings underscore the potential of TIL-based biomarkers, while also highlighting the need for more specific immune subsets that can better capture clinically relevant immune surveillance in the post-NACRT setting.

CD103 (ITGAE) marks a subset of T_RM_-like CD8⁺ T cells with durable cytotoxic function and epithelial retention via E-cadherin binding [7]. High CD103⁺CD8⁺ T-cell infiltration has been associated with favorable outcomes in lung and melanoma [8,9], and prognostic relevance has also been reported in surgically treated colorectal cancer [10–12].

Whether this prognostic significance is maintained after NACRT remains unknown. NACRT profoundly remodels the tumor microenvironment, and it is unclear whether CD103⁺CD8⁺ T cells in post-treatment specimens are treatment-induced or represent pre-existing populations. In rectal cancer after NACRT, neither the prognostic significance nor the origins of CD103⁺CD8⁺ T cells have been systematically investigated.

In this study, we applied Cu-Cyto, a deep learning–based imaging cytometry platform previously developed and validated [13], to quantify CD103⁺CD8⁺ T cells in post-NACRT rectal cancer specimens. This automated approach enables objective, single-cell–level identification and compartment-specific phenotyping across large tissue areas, minimizing observer-dependent bias. We evaluated the prognostic significance of CD103⁺CD8⁺ T-cell density in stromal and intratumoral compartments, assessed treatment-induced dynamics using paired biopsy–resection specimens, and explored their potential to stratify patients for adjuvant chemotherapy.

## Materials and Methods

### Patient cohort

Between January 2005 and December 2016, 47 patients with locally advanced rectal adenocarcinoma underwent NACRT followed by radical resection at Kobe University Hospital. Patients were eligible if they had pathologically confirmed adenocarcinoma, clinical stage T3–4 and/or node-positive disease without distant metastasis (M0) per the UICC 7th edition [14], and adequate tissue specimens available for analysis. Of these, 7 patients who achieved pathological complete response (grade 3 per the Japanese Classification of Colorectal Carcinoma [15]) were excluded. The remaining 40 patients constituted the study cohort (Fig. S1).

### Treatment

NACRT comprised pelvic radiotherapy (45–50.4 Gy in 25–28 fractions) combined with oral 5-fluorouracil–based chemotherapy, as previously described [16]. Radical resection was performed 6–8 weeks after NACRT completion via open or laparoscopic approaches, following total mesorectal excision principles. Lateral pelvic lymph node dissection was added selectively when pretreatment imaging suggested lateral lymph node metastasis. AC was administered to patients without contraindications related to comorbidities or performance status.

### Pathological assessment of tumor response

Pathological tumor response was graded by experienced pathologists per the Japanese Classification of Colorectal Carcinoma [15]: Grade 0 (no effect); Grade 1a (<1/3 of lesion affected); Grade 1b (≥1/3 to <2/3 affected); Grade 2 (≥2/3 affected with residual viable tumor); and Grade 3 (complete response, no viable tumor). Patients with Grade 3 were excluded, as residual tumor tissue was required for T-cell quantification.

### Immunohistochemistry and image acquisition

Formalin-fixed, paraffin-embedded tumor sections (4 μm) were stained with monoclonal antibodies against CD8 (Nichirei Biosciences, Tokyo, Japan) and CD103 (Eurobio Scientific, Les Ulis, France), with hematoxylin counterstaining. Stained slides were digitized at ×20 magnification using a NanoZoomer 2.0 HT whole-slide scanner (Hamamatsu Photonics, Hamamatsu, Japan).

### Compartmental segmentation

Tumor regions were classified into stromal and intratumoral compartments per the International TILs Working Group recommendations [17]. Stromal regions were defined as connective tissue surrounding tumor nests; intratumoral regions were defined as tumor cell nests. Areas of necrosis or prominent artifacts were excluded. Representative examples are shown in Figure 1.

**Figure 1.**
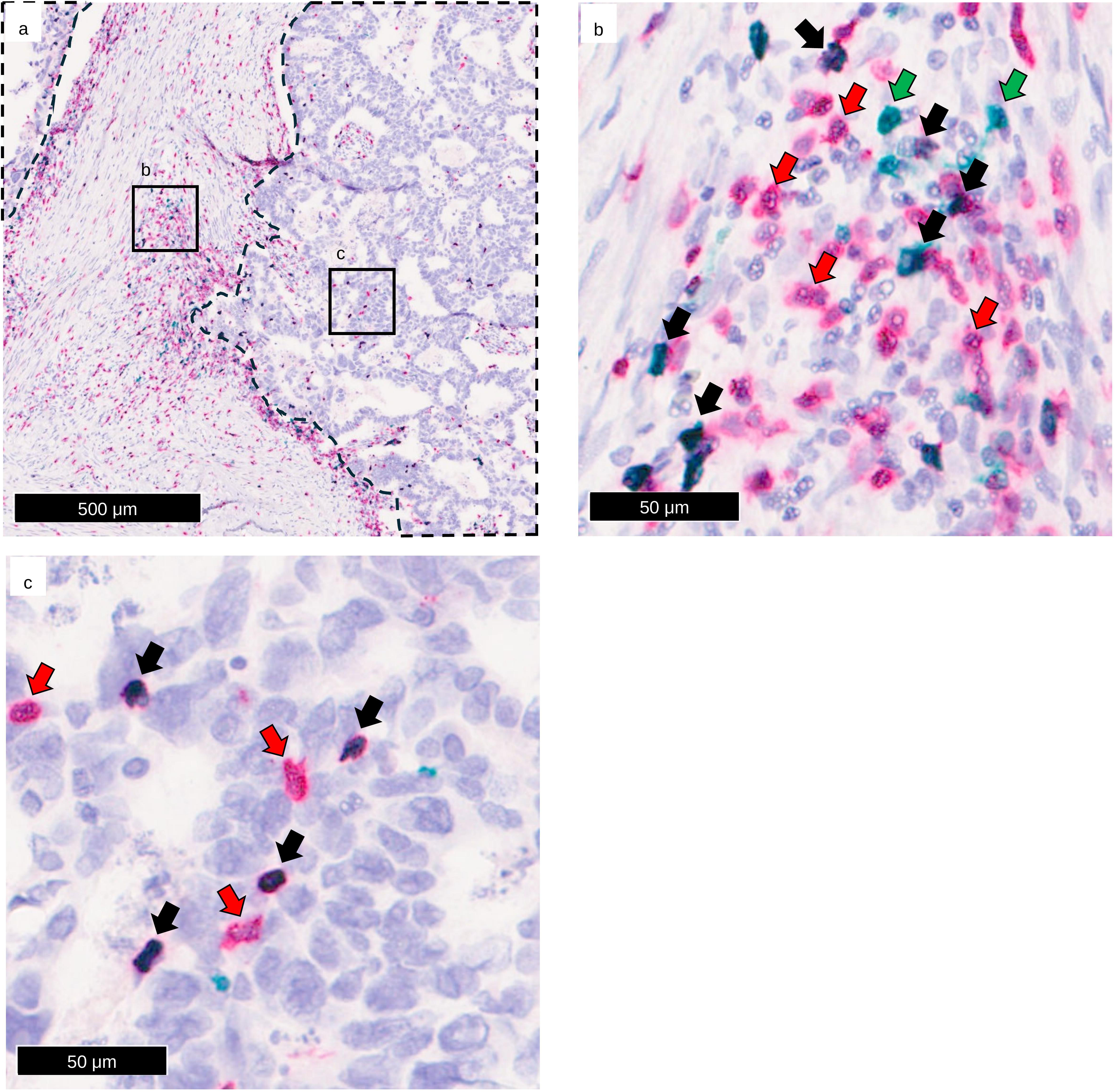
Compartmentalized immunohistochemical identification of CD8⁺ and CD103⁺CD8⁺ T cells in rectal cancer after neoadjuvant chemoradiotherapy (NACRT). (a) Representative histopathological section of rectal adenocarcinoma after NACRT, stained with hematoxylin, anti-CD8 (red), and anti-CD103 (green). The tumor–stroma interface is indicated by a black dotted line, illustrating the compartmental separation used for subsequent quantitative analyses. Scale bar = 500 μm. (b, c) Higher-magnification views of the boxed area in (a), showing representative examples of the stromal compartment (b) and the intratumoral compartment (c). Scale bar = 50 μm. CD103⁻CD8⁺ T cells are shown in red (red arrows), CD103⁺CD8⁻ cells in green (green arrows), and CD103⁺CD8⁺ double-positive T cells in black (black arrows).

### Quantification of immune cells using image cytometry (Cu-Cyto)

Immune cell densities were quantified using Cu-Cyto, a deep learning–based imaging cytometry platform previously developed and validated [13]. Cu-Cyto performs multicellular identification, simultaneously classifying multiple cell types within a single image—including both immunostained cells and morphologically defined populations that are not distinguishable by immunostaining. The training data, validation strategy, and performance benchmarks supporting its accuracy have been reported previously [13].

For each case, at least 10 image patches (146 μm × 146 μm) were randomly sampled from each compartment. Mean densities of CD8⁺ and CD103⁺CD8⁺ T cells (cells/mm²) were calculated per compartment, along with the proportion of CD103⁺ cells among total CD8⁺ T cells. Patches containing necrosis, tissue folds, or artifacts were excluded.

### Paired biopsy and resection specimens

Paired pretreatment biopsy and post-NACRT resection specimens were available in 9 patients. Cases were included only when at least 10 evaluable image patches were obtainable from both stromal and intratumoral compartments in each specimen, ensuring robust quantification. Immune cell densities were quantified using the Cu-Cyto pipeline described above, enabling direct intra-patient comparison of treatment-induced immune changes.

### Statistical analysis

The primary endpoints were relapse-free survival (RFS) and overall survival (OS). RFS was defined as the time from surgery to first recurrence or death from any cause; OS as the time from surgery to death from any cause. Patients without events were censored at last follow-up, summarized by the reverse Kaplan–Meier method.

Data normality was assessed by the Shapiro-Wilk test. Between-group comparisons used Student’s *t*-test (two groups) or the Kruskal-Wallis test (three or more groups). Optimal cutoff values for immune metrics were determined by ROC curve analysis for RFS (Fig. S2); as cutoffs were derived and tested in the same cohort, sensitivity analyses across clinical subgroups were performed to address potential optimism bias. Survival was estimated by the Kaplan–Meier method and compared by log-rank test. Variables significant at *p* < 0.05 on univariate analysis were entered into multivariate Cox models. Correlated immune markers were assessed in separate multivariate models to minimize collinearity. No stepwise variable selection was performed. The proportional hazards assumption was verified by log-minus-log plots. Results are expressed as hazard ratios (HRs) with 95% confidence intervals (CIs); *p* < 0.05 (two-sided) was considered significant.

Stromal and intratumoral compartments were analyzed separately. Supportive validation using manual pathologist counts and proportion-based analyses are presented in Figs. S3–S5. All analyses used SPSS Statistics version 29 (IBM Corp., Armonk, NY).

## Results

### Patient characteristics

The study cohort comprised 40 patients; clinicopathological characteristics are summarized in Tables S1 and S2. The median age was 67.5 years (range, 39–88), and 29 patients (72.5%) were male. Clinical T stage was T3 in 32 patients (80.0%) and T4 in 8 (20.0%); 22 patients (55.0%) had clinically node-positive disease.

Pathological tumor response was evaluable in 38 patients (2 excluded for missing data). Among these, 15 (39.5%) showed grade 2 response, 10 (25.0%) grade 1b, and 13 (32.5%) grade 1a. Twelve patients (30.0%) were classified as ypStage III and 28 (70.0%) as ypStage < III. AC was administered to 18 patients (45.0%). Lateral pelvic lymph node dissection had been performed in 23 patients (57.5%).

Median follow-up was 52 months (range, 5–143). Disease recurrence occurred in 17 patients (42.5%) and 13 (32.5%) died.

### Distribution of CD8⁺ and CD103⁺CD8⁺ T cells in stromal and intratumoral compartments

CD8⁺ and CD103⁺CD8⁺ T-cell densities were quantified in stromal and intratumoral compartments using Cu-Cyto (Figure 1). Stromal CD8⁺ density exceeded intratumoral density (1139 ± 80 vs. 278 ± 51 cells/mm², *p* < 0.001; Fig. 2a), as did stromal CD103⁺CD8⁺ density (100 ± 20 vs. 46 ± 13 cells/mm², *p* = 0.023; Fig. 2b). Conversely, the proportion of CD103⁺ cells among CD8⁺ T cells was higher in the intratumoral than stromal compartment (14.4% ± 2.2% vs. 8.8% ± 1.5%, *p* = 0.042; Fig. 2c).

**Figure 2.**
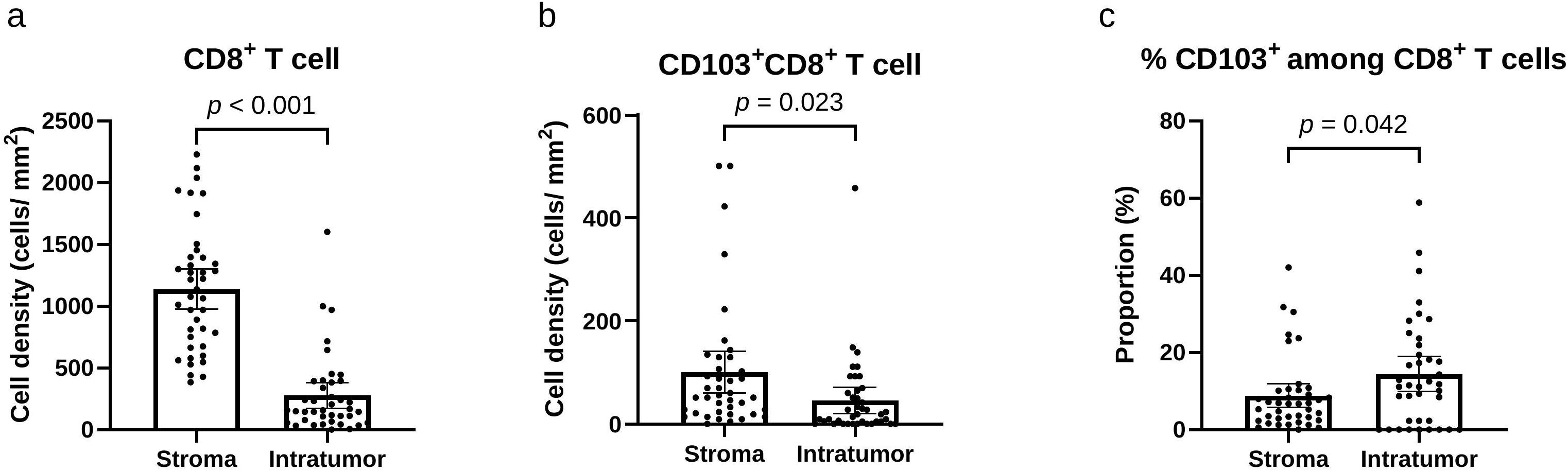
Differential distribution of CD8⁺ and CD103⁺CD8⁺ T cells between stromal and intratumoral compartments after neoadjuvant chemoradiotherapy (NACRT). (a) Comparison of CD8⁺ T-cell densities between the stromal and intratumoral compartments. (b) Comparison of CD103⁺CD8⁺ T-cell densities between the stromal and intratumoral compartments. (c) Proportion of CD103⁺ cells among total CD8⁺ T cells in each compartment. Although the absolute densities of both CD8⁺ and CD103⁺CD8⁺ T cells were higher in the stromal compartment, the intratumoral compartment showed a higher proportion of CD103⁺ cells among CD8⁺ T cells. Data are presented as mean ± SD. Statistical significance was assessed using the Wilcoxon signed-rank test. Each dot represents one patient (*n* = 40).

### Association between T-cell densities and pathological tumor response

We next examined the association between pathological tumor response grade and T-cell densities (Fig. 3). Stromal CD8⁺ density correlated positively with response grade (*p* = 0.037; Fig. 3a), whereas intratumoral CD8⁺ density did not (Fig. 3b). Neither stromal nor intratumoral CD103⁺CD8⁺ density was associated with response grade (Fig. 3c, d).

**Figure 3.**
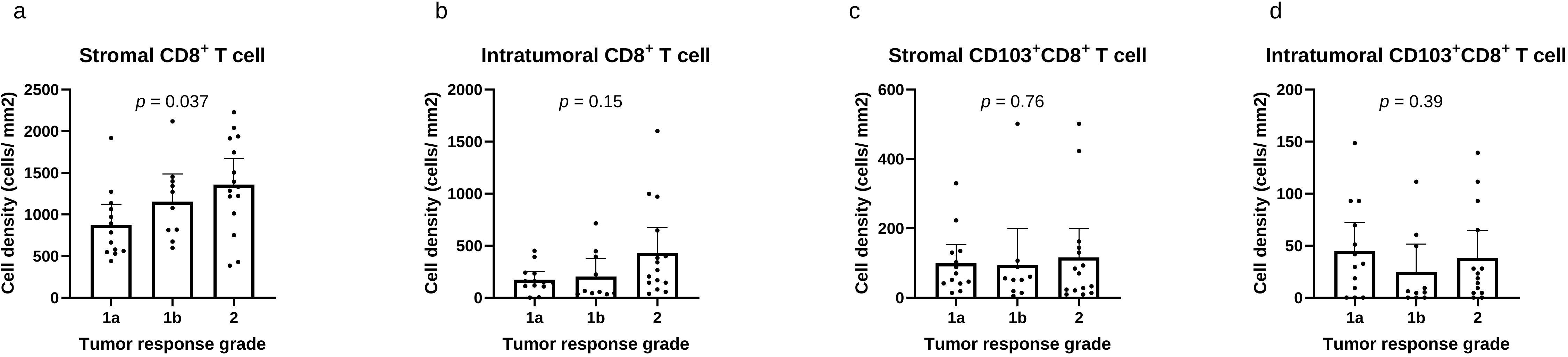
Association between T-cell densities and pathological tumor response grade after neoadjuvant chemoradiotherapy (NACRT). Comparison of T-cell subset densities across pathological tumor response grades defined by the Japanese Classification of Colorectal Carcinoma: (a) stromal CD8⁺ T cells, (b) intratumoral CD8⁺ T cells, (c) stromal CD103⁺CD8⁺ T cells, and (d) intratumoral CD103⁺CD8⁺ T cells (*n* = 38; 2 patients excluded for missing tumor response grade data). Data are presented as mean ± SD. Statistical significance was assessed using the Kruskal–Wallis test. A significant association with pathological response grade was observed only for stromal CD8⁺ T cells (*p* < 0.05).

### Dynamic Changes in CD8⁺ and CD103⁺CD8⁺ T cells before and after NACRT

To investigate immunological alterations induced by NACRT, we compared CD8⁺ and CD103⁺CD8⁺ T-cell densities between paired pretreatment biopsy and post-NACRT resection specimens. Of the available paired cases, 9 met the criterion of at least 10 evaluable patches per compartment in both specimens, ensuring quantification robustness (Fig. 4). Stromal CD8⁺ density increased significantly after NACRT (741.3 ± 228.1 to 1405.1 ± 147.6 cells/mm²; *p* = 0.008; Fig. 4a), whereas intratumoral CD8⁺ density showed a non-significant trend toward increase (106.8 ± 33.6 to 277.0 ± 102.7 cells/mm²; *p* = 0.098; Fig. 4b). In contrast, neither stromal (114.0 ± 42.2 to 114.5 ± 50.1 cells/mm²; *p* = 0.999; Fig. 4c) nor intratumoral (34.0 ± 13.4 to 43.7 ± 14.0 cells/mm²; *p* = 0.496; Fig. 4d) CD103⁺CD8⁺ density changed significantly with treatment.

**Figure 4.**
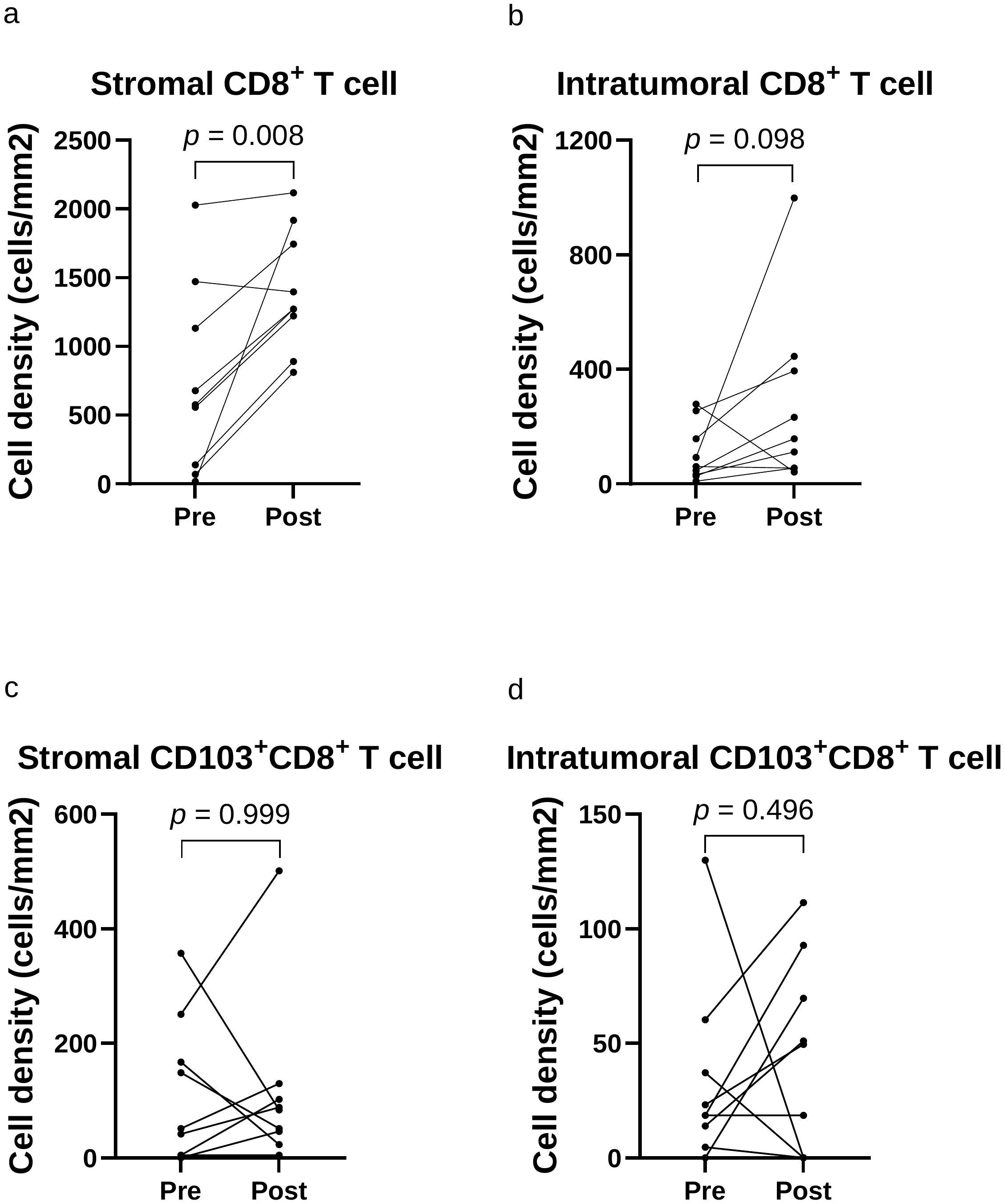
Dynamic changes in CD8⁺ and CD103⁺CD8⁺ T-cell densities before and after neoadjuvant chemoradiotherapy (NACRT). Comparison of CD8⁺ and CD103⁺CD8⁺ T-cell densities between paired pretreatment biopsy and post-NACRT resection specimens (*n* = 9) in the stromal and intratumoral compartments. Each line represents an individual patient. T-cell densities were quantified using the Cu-Cyto platform; only cases with at least 10 evaluable patches per compartment in both biopsy and resection specimens were included to ensure robust quantification. Statistical significance was assessed using the Wilcoxon signed-rank test. (a) Stromal CD8⁺ T-cell density (*p* = 0.008). (b) Intratumoral CD8⁺ T-cell density (*p* = 0.098). (c) Stromal CD103⁺CD8⁺ T-cell density (*p* = 0.999). (d) Intratumoral CD103⁺CD8⁺ T-cell density (*p* = 0.496).

### Prognostic impact of CD8⁺ T-cell density

We next assessed the prognostic significance of CD8⁺ density for RFS and OS (Fig. S3). High stromal CD8⁺ density was associated with significantly better RFS (*p* < 0.001; Fig. S3a), as was high intratumoral CD8⁺ density (*p* = 0.029; Fig. S3b). Neither compartment showed a significant association with OS (*p* = 0.135 and *p* = 0.110, respectively; Fig. S3c, d), though patients with higher densities tended toward better survival.

### Stromal CD103⁺CD8⁺ T-cell density and patient survival

We next examined the prognostic significance of CD103⁺CD8⁺ density for RFS and OS (Fig. 5). High stromal CD103⁺CD8⁺ density was associated with significantly better RFS (*p* < 0.001; Fig. 5a) and OS (*p* = 0.016; Fig. 5c), whereas intratumoral CD103⁺CD8⁺ density showed no significant association with either endpoint (Fig. 5b, d).

**Figure 5.**
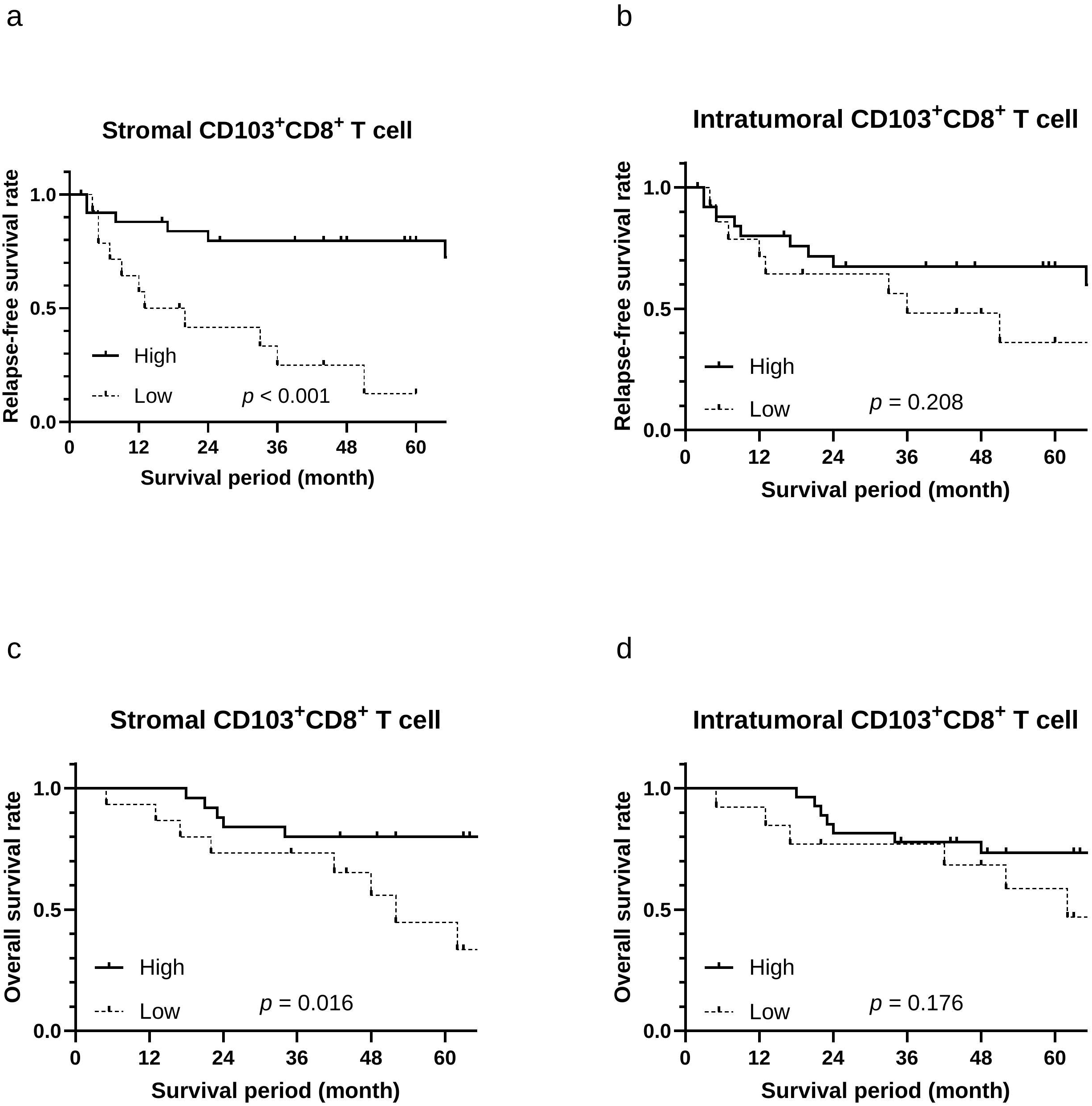
Survival analyses according to stromal and intratumoral CD103⁺CD8⁺ T-cell density after neoadjuvant chemoradiotherapy (NACRT). Kaplan–Meier curves for relapse-free survival (RFS) and overall survival (OS) stratified by CD103⁺CD8⁺ T-cell density in each compartment: (a) RFS according to stromal CD103⁺CD8⁺ T-cell density, (b) RFS according to intratumoral CD103⁺CD8⁺ T-cell density, (c) OS according to stromal CD103⁺CD8⁺ T-cell density, and (d) OS according to intratumoral CD103⁺CD8⁺ T-cell density. High stromal CD103⁺CD8⁺ T-cell density was significantly associated with both improved RFS (*p* < 0.001) and OS (*p* = 0.016). In contrast, CD103⁺CD8⁺ T-cell density in the intratumoral compartment showed no significant association with either endpoint. Patients were dichotomized into high- and low-density groups based on optimal cut-off values. Statistical significance was determined using the log-rank test.

Cox regression analyses are summarized in Table 1. On univariate analysis, stromal CD103⁺CD8⁺ density (HR, 0.168; 95% CI, 0.057–0.494; *p* = 0.001), stromal CD8⁺ density (HR, 0.176; 95% CI, 0.040–0.775; *p* = 0.022), and intratumoral CD8⁺ density (HR, 0.356; 95% CI, 0.134–0.943; *p* = 0.038) were each associated with RFS. On multivariate analysis adjusted for ypStage, stromal CD103⁺CD8⁺ density (HR, 0.188; 95% CI, 0.062–0.570; *p* = 0.003) and stromal CD8⁺ density (HR, 0.158; 95% CI, 0.035–0.711; *p* = 0.016) retained independent significance for RFS, whereas intratumoral CD8⁺ density did not (HR, 0.398; *p* = 0.069).

**Table 1.** Univariate and multivariate Cox proportional hazards regression analyses for relapse-free survival (RFS) and overall survival (OS). Hazard ratios (HRs) and 95% confidence intervals (CIs) for clinicopathological variables and immune cell densities in patients with rectal cancer after neoadjuvant chemoradiotherapy (NACRT). Stromal and intratumoral CD8⁺ and CD103⁺CD8⁺ T-cell densities were dichotomized into high and low groups according to receiver operating characteristic (ROC)-derived cut-off values. Bold values indicate statistical significance (*p* < 0.05).

|  | Univariate | Univariate | Multivariate-1 | Multivariate-1 | Multivariate-2 | Multivariate-2 | Multivariate-3 | Multivariate-3 | Multivariate-3 |  |
| --- | --- | --- | --- | --- | --- | --- | --- | --- | --- | --- |
| Relapse-free survival | HR (95% CI) | p value | HR (95% CI) | p value | HR (95% CI) | p value | p value | HR (95% CI) | p value | p value |
| Age (≥ 78/ < 78 y.o.) | 2.749 (0.786-9.619) | 0.114 |  |  |  |  |  |  |  |  |
| Sex (M/ F) | 0.495 (0.142-1.730) | 0.271 |  |  |  |  |  |  |  |  |
| Operative time (≥ 769/ < 769min) | 1.544 (0.502-4.753) | 0.449 |  |  |  |  |  |  |  |  |
| Blood loss (≥ 315/ < 315mL) | 2.147 (0.791-5.829) | 0.134 |  |  |  |  |  |  |  |  |
| CEA after NACRT (≥ 5/ < 5 ng/ml) | 0.624 (0.199-1.951) | 0.417 |  |  |  |  |  |  |  |  |
| CA19-9 after NACRT (≥ 37/ < 37 U/ml) | 1.379 (0.476-3.992) | 0.554 |  |  |  |  |  |  |  |  |
| ypStage (≥ III/ < III) | 2.737 (1.048-7.147) | 0.040 | 3.110 (1.160-8.340) | 0.024 | 2.470 (0.927-6.581) | 0.071 | 0.071 | 2.032 (0.744-5.552) | 0.167 | 0.167 |
| Pathological tumor response grade (Grade ≥ 2/ < 2) | 0.577 (0.2-1.664) | 0.308 |  |  |  |  |  |  |  |  |
| Adjuvant chemotherapy (+/ -) | 0.666 (0.253-1.757) | 0.412 |  |  |  |  |  |  |  |  |
| Stromal CD8+ T-cell density (≥ 1278.2/ < 1278.2 cells/mm2) | 0.176 (0.040-0.775) | 0.022 | 0.158 (0.035-0.711) | 0.016 |  |  |  |  |  |  |
| Intratumoral CD8+ T-cell density (≥ 60.35/ < 60.35 cells/mm2) | 0.356 (0.134-0.943) | 0.038 |  |  | 0.398 (0.147-1.075) | 0.069 | 0.069 |  |  |  |
| Stromal CD103+CD8+ T-cell density (≥ 40.9/ < 40.9 cells/mm2) | 0.168 (0.057-0.494) | 0.001 |  |  |  |  |  | 0.188 (0.062-0.570) | 0.003 | 0.003 |
| Intratumoral CD103+CD8+ T-cell density (≥ 7.74/ < 7.74 cells/mm2) | 0.546 (0.210-1.425) | 0.217 |  |  |  |  |  |  |  |  |
|  | Univariate | Univariate | Multivariate | Multivariate |  |  |  |  |  |  |
| Overall survival | HR (95% CI) | p value | HR (95% CI) | p value |  |  |  |  |  |  |
| Age (≥ 75/ < 75 y.o.) | 5.049 (1.631-15.634) | 0.005 | 8.093 (2.297-28.522) | 0.001 |  |  |  |  |  |  |
| Sex (M/ F) | 0.199 (0.026-1.530) | 0.121 |  |  |  |  |  |  |  |  |
| Operative time (≥ 400/ <400 min) | 1.298 (0.357-4.720) | 0.692 |  |  |  |  |  |  |  |  |
| Blood loss (≥ 143/ <143 mL) | 1.853 (0.570-6.020) | 0.305 |  |  |  |  |  |  |  |  |
| CEA after NACRT (≥ 5/ < 5 ng/ml) | 0.406 (0.089-1.860) | 0.246 |  |  |  |  |  |  |  |  |
| CA19-9 after NACRT (≥ 37/ < 37 U/ml) | 2.205 (0.691-7.043) | 0.182 |  |  |  |  |  |  |  |  |
| ypStage (≥ III/ < III) | 2.292 (0.768-6.838) | 0.137 |  |  |  |  |  |  |  |  |
| Pathological tumor response grade (Grade ≥ 2/ < 2) | 0.304 (0.066-1.408) | 0.128 |  |  |  |  |  |  |  |  |
| Adjuvant chemotherapy (+/ -) | 0.925 (0.210-2.756) | 0.888 |  |  |  |  |  |  |  |  |
| Stromal CD8+ T-cell density (≥ 1107.2/ < 1107.2 cells/mm2) | 0.417 (0.128-1.360) | 0.147 |  |  |  |  |  |  |  |  |
| Intratumoral CD8+ T-cell density (≥ 547.8/ < 547.8 cells/mm2) | 0.037 (0.000-22.436) | 0.313 |  |  |  |  |  |  |  |  |
| Stromal CD103+CD8+ T-cell density (≥ 40.9/ < 40.9 cells/mm2) | 0.275 (0.089-0.848) | 0.025 | 0.184 (0.054-0.626) | 0.007 |  |  |  |  |  |  |
| Intratumoral CD103+CD8+ T-cell density (≥ 4.9/ < 4.9 cells/mm2) | 0.479 (0.161-1.426) | 0.186 |  |  |  |  |  |  |  |  |

For OS, stromal CD103⁺CD8⁺ density was the only immune variable that retained independent prognostic significance on multivariate analysis adjusted for age (HR, 0.184; 95% CI, 0.054–0.626; *p* = 0.007; Table 1).

### Adjuvant chemotherapy stratification according to stromal CD103⁺CD8⁺ T-cell density

We explored whether stromal CD103⁺CD8⁺ density could stratify the benefit of AC on RFS (Fig. 6). Stratification by stromal CD8⁺ density alone revealed no differential effect of AC in either subgroup (Fig. 6a, b). In contrast, among patients with low stromal CD103⁺CD8⁺ density, AC was associated with better RFS (5-year RFS, 25% vs. 0%; *p* = 0.049; Fig. 6c), whereas no such association was observed in the high-density subgroup (Fig. 6d). These findings are exploratory and require prospective validation.

**Figure 6.**
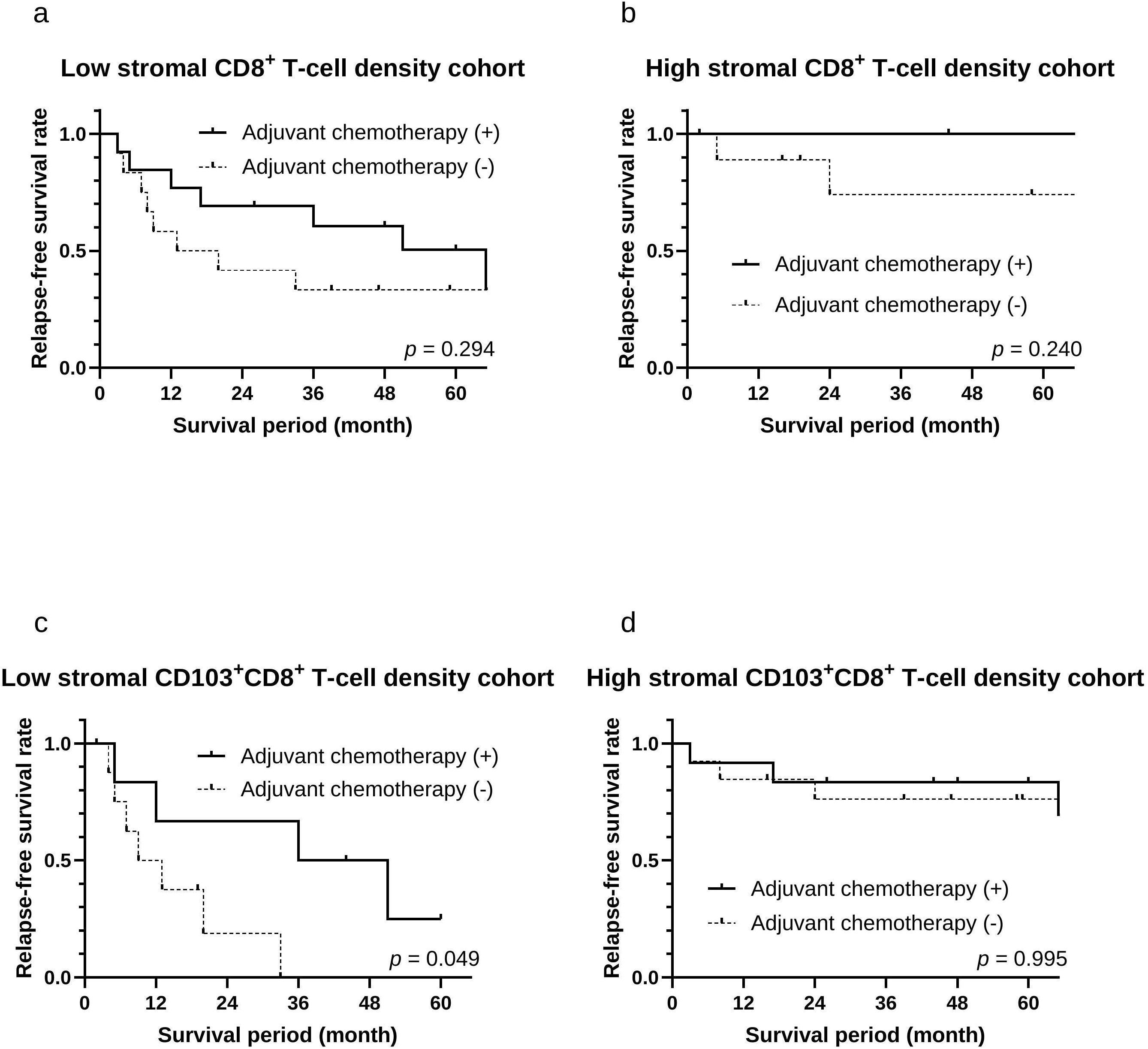
Exploratory analysis of relapse-free survival stratified by stromal T-cell density and receipt of adjuvant chemotherapy. Kaplan–Meier curves for relapse-free survival (RFS) stratified by stromal T-cell density (high vs. low) and receipt of adjuvant chemotherapy (AC). (a, b) Stratification by stromal CD8⁺ T-cell density: (a) low stromal CD8⁺ T-cell density (*n* = 25; AC, *n* = 13; no AC, *n* = 12; *p* = 0.294) and (b) high stromal CD8⁺ T-cell density (*n* = 15; AC, *n* = 5; no AC, *n* = 10; *p* = 0.240). No significant differential effect of AC on RFS was observed in either subgroup. (c, d) Stratification by stromal CD103⁺CD8⁺ T-cell density: (c) low stromal CD103⁺CD8⁺ T-cell density (*n* = 15; AC, *n* = 6; no AC, *n* = 9; *p* = 0.049) and (d) high stromal CD103⁺CD8⁺ T-cell density (*n* = 25; AC, *n* = 12; no AC, *n* = 13; *p* = 0.995). In the low CD103⁺CD8⁺ density group, AC was associated with a trend toward improved RFS (5-year RFS, 25% with AC vs. 0% without AC; *p* = 0.049), whereas no apparent benefit of AC was observed in the high CD103⁺CD8⁺ density group. These results should be interpreted as exploratory.

### Validation and sensitivity analyses

Manual pathologist counts yielded results consistent with Cu-Cyto quantification, with stromal CD103⁺CD8⁺ density remaining associated with favorable outcomes (Figs. S3, S4; Table S3). When the proportion of CD103⁺ among CD8⁺ T cells was used instead of absolute density, no significant survival association was observed (Fig. S5). Sensitivity analyses confirmed the prognostic stability of stromal CD103⁺CD8⁺ density across subgroups defined by AC receipt, ypStage, and follow-up ≥ 36 months (Table S4).

## Discussion

We evaluated the prognostic significance of CD8⁺ and CD103⁺CD8⁺ T cells in post-NACRT rectal cancer using Cu-Cyto, a deep learning-based imaging cytometry platform. Stromal CD103⁺CD8⁺ T-cell density emerged as an independent prognostic factor for both RFS and OS, and exploratory analyses suggested that AC may preferentially benefit patients with low stromal CD103⁺CD8⁺ density. To our knowledge, no previous study has systematically evaluated compartment-specific CD103⁺CD8⁺ T-cell density by immunohistochemistry in NACRT-treated rectal cancer.

CD8⁺ T cells are consistently associated with favorable prognosis in colorectal and other solid tumors [18], consistent with prior studies in rectal cancer after NACRT. In the present cohort, both stromal and intratumoral CD8⁺ density were associated with RFS, reinforcing the prognostic role of CD8⁺ T cells in the post-NACRT setting and supporting the validity of our quantification approach.

CD103 marks T_RM_-like CD8⁺ T cells with durable cytotoxic function and epithelial retention via E-cadherin binding [7]. High CD103⁺CD8⁺ T-cell infiltration has been associated with favorable outcomes and immunotherapy responsiveness in lung and melanoma [8,9], and prognostic relevance has also been reported in surgically treated colorectal cancer [10–12]. The present findings extend T_RM_ biology into the post-NACRT setting and identify the stromal compartment as the prognostically informative niche.

Functional heterogeneity within the CD103⁺CD8⁺ T-cell compartment has been increasingly recognized [11,19]. Co-expression of CD69, CD49a, and CD39 defines biologically distinct subsets: CD69 marks tissue-retentive cells associated with favorable outcomes [11,12], whereas CD49a and CD39 identify tumor-reactive, cytotoxic populations with independent prognostic value [12,19,20,21]. As the present study relied on CD103 alone to define T_RM_-like T cells, these phenotypic distinctions were not captured. Whether the differential distribution of such subsets between stromal and intratumoral compartments underlies the compartment-specific prognostic effect observed here remains to be determined.

A key question is whether CD103⁺CD8⁺ T cells in post-NACRT specimens are treatment-induced or pre-existing. If treatment-induced, their density would be expected to correlate with pathological response and to increase alongside total CD8⁺ T cells after NACRT. Neither was observed. Stromal CD103⁺CD8⁺ density showed no association with pathological response grade (*p* = 0.76), in contrast to stromal CD8⁺ density (*p* = 0.037). Paired pre-/post-NACRT analysis revealed a selective non-expansion of the CD103⁺ subset: stromal CD8⁺ density increased significantly (*p* = 0.008) while stromal CD103⁺CD8⁺ density remained unchanged (*p* = 0.999), and a parallel pattern was observed in the intratumoral compartment. This dissociation between total CD8⁺ expansion and CD103⁺CD8⁺ stability argues against NACRT-driven generation of the CD103⁺CD8⁺ population and is more consistent with a pre-existing tissue-resident pool, although formal validation would require TCR repertoire or lineage-tracing analyses beyond the scope of the present work.

In cervical cancer, Komdeur et al. reported that the prognostic impact was confined to the radiotherapy-treated subgroup [22], suggesting radiation may render T_RM_-like immunity clinically decisive. Our data support a different interpretation: stromal CD103⁺CD8⁺ density was neither induced by NACRT nor correlated with pathological response, yet independently predicted RFS and OS. Combined with consistent prognostic findings in surgically resected colorectal cancer cohorts, this suggests that CD103⁺CD8⁺ T cells, as T_RM_-like T cells, exhibit intrinsic immunological capacity that remains prognostically informative across both upfront-surgery and post-NACRT contexts.

The dissociation between total CD8⁺ and CD103⁺CD8⁺ T cells underscores that the latter captures prognostic information distinct from total CD8⁺ assessment, reflecting intrinsic immune competence rather than a treatment-induced effect. This stability across NACRT raises the possibility that pretreatment biopsy profiling may provide equivalent prognostic information.

Stromal CD103⁺CD8⁺ density has two potential clinical applications. First, it may stratify recurrence risk after NACRT: patients with high density had markedly favorable outcomes, whereas those with low density faced substantially higher recurrence risk, consistent with other tumor types [8,23]. Second, exploratory data suggest AC may benefit predominantly low-density patients; if validated, this could refine postoperative decisions by sparing high-density patients from unnecessary toxicity [24].

Recent phase II and randomized trials have shown that adding ICB to NACRT improves pathological complete response rates in microsatellite-stable LARC [25–28], although whether this translates into a survival benefit remains unclear. Several trials administer ICB sequentially after CRT and prior to surgery, positioning the resected specimen as a directly informative timepoint for immune assessment. In several solid tumors, high intratumoral CD103⁺CD8⁺ T-cell density has been associated with favorable ICB outcomes [29–31], suggesting that patients with high density may be particularly suited to ICB-based regimens. Because CD103⁺CD8⁺ T-cell density appears to reflect pre-existing rather than treatment-induced immunity, pretreatment biopsy specimens may capture equivalent prognostic information—providing a practical basis for patient stratification before therapy begins. Prospective studies in ICB-treated cohorts are needed to determine whether pretreatment CD103⁺CD8⁺ density can guide treatment selection.

This study has several limitations. First, the single-institution retrospective design and small cohort (n = 40; 17 recurrence events for RFS and 13 deaths for OS) limit generalizability; the limited number of events relative to covariates in multivariate models falls below the conventional benchmark of 10 events per variable and increases the risk of overfitting; multivariate results should therefore be interpreted as hypothesis-generating rather than confirmatory. Second, exclusion of pCR patients—who typically have favorable outcomes—may have shifted the cohort toward higher risk. Third, heterogeneity in radiation dose and chemotherapy regimens may have influenced immune profiles. Fourth, the paired analysis was limited to 9 patients, though its findings were consistent with the lack of association between CD103⁺CD8⁺ density and pathological response in the full cohort. Fifth, the AC stratification analysis was exploratory and subject to non-randomized treatment allocation. Because AC was not randomly assigned, prognostic factors that also influenced treatment decisions—particularly ypStage and pathological tumor response grade—may confound the observed association within the low-density subgroup. Sixth, while our data suggest the prognostic equivalence of pretreatment biopsy specimens, this was not directly tested in a prospective cohort; dedicated prospective studies with systematic pretreatment immune assessment are required to establish clinical utility.

## Conclusions

Stromal CD103⁺CD8⁺ T-cell density is a robust independent prognostic biomarker in rectal cancer after NACRT. High density marked patients with favorable survival; low density identified those at high recurrence risk. Exploratory analyses further suggest AC may preferentially benefit low-density patients. Given that CD103⁺CD8⁺ T-cell density appears to reflect pre-existing rather than treatment-induced immunity, pretreatment biopsy assessment may offer equivalent prognostic information, opening the possibility of immune-based patient stratification before treatment initiation. Multi-institutional prospective validation is needed to confirm clinical utility and to determine whether pretreatment CD103⁺CD8⁺ density can guide individualized treatment decisions, including the selection of ICB-based strategies.

## Abbreviations

AC: adjuvant chemotherapy
Cu-Cyto: deep learning–based imaging cytometry platform
LARC: locally advanced rectal cancer
NACRT: neoadjuvant chemoradiotherapy
OS: overall survival
RFS: relapse-free survival
TIL: tumor-infiltrating lymphocyte
T_RM_: tissue-resident memory

## Acknowledgements

The authors thank the staff of the Department of Surgery, Kobe University Hospital, for their clinical support. A large language model (Claude, Anthropic) assisted with manuscript drafting and language editing; all outputs were critically reviewed and edited by the authors, who take full responsibility for the content.

## Author Contributions

Conceptualization, T.A., K.Y. and S.T.; methodology, T.A., K.Y. and T.N.; software, T.N., Y.U., T.Y. and T.Y.; validation, T.A., T.N., Y.U., S.M., Y.A., M.A., T.T., Y.O., R.K., T.T., H.K., T.T. and M.I.; formal analysis, T.A.; investigation, T.A.; resources, All authors; data curation, All authors; writing-original draft preparation, T.A; writing-review and editing, All authors; visualization, T.A. and T.N.; supervision, K.Y., T.M., M.F. H.Y., R.S., T.F. and Y.K.; project administration, K.Y.; funding acquisition, T.A., T.N., H.K., K.Y., T.M. and Y.K. All authors have read and agreed to the published version of the manuscript.

## Ethics Statement

The study was conducted in accordance with the Declaration of Helsinki and approved by the Institutional Review Board and Ethics Committee of Kobe University (Approval No: B190154, approved on 17 Sep 2019). Patient informed consent was waived by the Institutional Review Board and Ethics Committee of Kobe University due to the retrospective nature of the study.

## Data Availability Statement

Data are available from the corresponding author upon reasonable request. Data sharing will require additional ethical board approval.

## Supplementary Information

Fig. S1: Study design and patient flow

Fig. S2: ROC curve analyses for determination of optimal cut-off values

Fig. S3: Prognostic significance of CD8⁺ T-cell density (Cu-Cyto quantification and manual pathologist quantification)

Fig. S4: Kaplan–Meier survival curves based on manual pathologist quantification (CD103⁺CD8⁺ T cells)

Fig. S5: Kaplan–Meier survival curves based on proportion of CD103⁺ among CD8⁺ T cells

Table S1: Clinicopathological characteristics stratified by stromal CD103⁺CD8⁺ T-cell density

Table S2: Patient clinicopathological characteristics

Table S3: Univariate and multivariate Cox proportional hazards regression models for relapse-free survival (RFS) and overall survival (OS) based on manual pathologist quantification

Table S4: Sensitivity analyses for relapse-free survival (RFS)

## References

1. Guillem JG, Chessin DB, Cohen AM, et al. Long-term oncologic outcome following preoperative combined modality therapy and total mesorectal excision of locally advanced rectal cancer. Ann Surg 241(5), 829–838 (2005). doi:10.1097/01.sla.0000161980.46459.96

2. Sauer R, Becker H, Hohenberger W, et al. Preoperative versus Postoperative Chemoradiotherapy for Rectal Cancer. N Engl J Med 351(17), 1731–1740 (2004). doi:10.1056/NEJMoa040694

3. Yasuda K, Nirei T, Sunami E, et al. Density of CD4(+) and CD8(+) T Lymphocytes in Biopsy Samples Can Be a Predictor of Pathological Response to Chemoradiotherapy (CRT) for Rectal Cancer. Radiat Oncol 6, 49 (2011). doi:10.1186/1748-717X-6-49

4. Anitei M-G, Zeitoun G, Mlecnik B, et al. Prognostic and Predictive Values of the Immunoscore in Patients with Rectal Cancer. Clin Cancer Res 20(7), 1891–1899 (2014). doi:10.1158/1078-0432.CCR-13-2830

5. Ogura A, Akiyoshi T, Yamamoto N, et al. Pattern of Programmed Cell Death-Ligand 1 Expression and CD8-Positive T-Cell Infiltration before and after Chemoradiotherapy in Rectal Cancer. Eur J Cancer 91, 11–20 (2018). doi:10.1016/j.ejca.2017.12.005

6. Wirta E-V, Elomaa H, Ahtiainen M, et al. The Impact of Preoperative Treatments on the Immune Environment of Rectal Cancer. APMIS 132(12), 1046–1060 (2024). doi:10.1111/apm.13467

7. Okła K, Farber DL, Zou W. Tissue-Resident Memory T Cells in Tumor Immunity and Immunotherapy. J Exp Med 218(4), e20201605 (2021). doi:10.1084/jem.20201605

8. Djenidi F, Adam J, Goubar A, et al. CD8+CD103+ Tumor–Infiltrating Lymphocytes Are Tumor-Specific Tissue-Resident Memory T Cells and a Prognostic Factor for Survival in Lung Cancer Patients. J Immunol 194(7), 3475–3486 (2015). doi:10.4049/jimmunol.1402711

9. Edwards J, Wilmott JS, Madore J, et al. CD103+ Tumor-Resident CD8+ T Cells Are Associated with Improved Survival in Immunotherapy-Naïve Melanoma Patients and Expand Significantly During Anti–PD-1 Treatment. Clin Cancer Res 24(13), 3036–3045 (2018). doi:10.1158/1078-0432.CCR-17-2257

10. Liu S, Wang P, Wang P, et al. Tissue-resident memory CD103+CD8+ T cells in colorectal cancer: its implication as a prognostic and predictive liver metastasis biomarker. Cancer Immunol Immunother 73(9), 176 (2024). doi:10.1007/s00262-024-03709-2

11. Wu ZX, Da TT, Huang C, et al. CD69+CD103+CD8+ tissue-resident memory T cells possess stronger anti-tumor activity and predict better prognosis in colorectal cancer. Cell Commun Signal 22(1), 608 (2024). doi:10.1186/s12964-024-01990-3

12. Talhouni S, Fadhil W, Mongan NP, et al. Activated tissue resident memory T-cells (CD8+CD103+CD39+) uniquely predict survival in left sided “immune-hot” colorectal cancers. Front Immunol 14, 1057292 (2023). doi:10.3389/fimmu.2023.1057292

13. Abe T, Yamashita K, Nagasaka T, et al. Deep learning-based image cytometry using a bit-pattern kernel-filtering algorithm to avoid multi-counted cell determination. Anticancer Res 43(8), 3755–3761 (2023). doi:10.21873/anticanres.16560

14. Sobin LH, Gospodarowicz MK, Wittekind C. TNM Classification of Malignant Tumours, 7th edn. (Wiley-Blackwell: Chichester, UK, 2009).

15. Japanese Society for Cancer of the Colon and Rectum. Japanese Classification of Colorectal, Appendiceal, and Anal Carcinoma, 3rd English edn. (Kanehara: Tokyo, Japan, 2019).

16. Matsuda T, Sumi Y, Yamashita K, et al. Outcomes and Prognostic Factors of Selective Lateral Pelvic Lymph Node Dissection with Preoperative Chemoradiotherapy for Locally Advanced Rectal Cancer. Int J Colorectal Dis 33(4), 367–374 (2018). doi:10.1007/s00384-018-2974-1

17. Salgado R, Denkert C, Demaria S, et al. The Evaluation of Tumor-Infiltrating Lymphocytes (TILs) in Breast Cancer: Recommendations by an International TILs Working Group 2014. Ann Oncol 26(2), 259–271 (2015). doi:10.1093/annonc/mdu450

18. Fridman WH, Zitvogel L, Sautès-Fridman C, Kroemer G. The Immune Contexture in Cancer Prognosis and Treatment. Nat Rev Clin Oncol 14(12), 717–734 (2017). doi:10.1038/nrclinonc.2017.101

19. Ganesan A-P, Clarke J, Wood O, et al. Tissue-Resident Memory Features Are Linked to the Magnitude of Cytotoxic T Cell Responses in Human Lung Cancer. Nat Immunol 18(8), 940–950 (2017). doi:10.1038/ni.3775

20. Abdeljaoued S, Doussot A, Kroemer M, et al. Liver Metastases of Colorectal Cancer Contain Different Subsets of Tissue-Resident Memory CD8 T Cells Correlated with a Distinct Risk of Relapse Following Surgery. OncoImmunology 14(1), 2455176 (2025). doi:10.1080/2162402X.2025.2455176

21. Ali A, Bari MF, Arshad S, et al. Tissue-Resident Memory T-Cell Expressions and Their Prognostic Role in Head and Neck Squamous Cell Carcinoma: A Systematic Review and Meta-Analysis. BMC Cancer 25(1), 356 (2025). doi:10.1186/s12885-025-13764-2

22. Komdeur FL, Prins TM, van de Wall S, et al. CD103+ Tumor-Infiltrating Lymphocytes Are Tumor-Reactive Intraepithelial CD8+ T Cells Associated with Prognostic Benefit and Therapy Response in Cervical Cancer. OncoImmunology 6(9), e1338230 (2017). doi:10.1080/2162402X.2017.1338230

23. Savas P, Virassamy B, Ye C, et al. Single-Cell Profiling of Breast Cancer T Cells Reveals a Tissue-Resident Memory Subset Associated with Improved Prognosis. Nat Med 24(7), 986–993 (2018). doi:10.1038/s41591-018-0078-7

24. Breugom AJ, Swets M, Bosset J-F, et al. Adjuvant Chemotherapy after Preoperative (Chemo)radiotherapy and Surgery for Patients with Rectal Cancer: A Systematic Review and Meta-Analysis of Individual Patient Data. Lancet Oncol 16(2), 200–207 (2015). doi:10.1016/S1470-2045(14)71199-4

25. Bando H, Tsukada Y, Inamori K, et al. Preoperative Chemoradiotherapy plus Nivolumab before Surgery in Patients with Microsatellite Stable and Microsatellite Instability–High Locally Advanced Rectal Cancer. Clin Cancer Res 28(6), 1136–1146 (2022). doi:10.1158/1078-0432.CCR-21-3213

26. Yang Z, Gao J, Zheng J, et al. Efficacy and safety of PD-1 blockade plus long-course chemoradiotherapy in locally advanced rectal cancer (NECTAR): a multi-center phase 2 study. Signal Transduct Target Ther 9(1), 56 (2024). doi:10.1038/s41392-024-01762-y

27. Hou Z, Liao L, Xiao W, et al. Neoadjuvant chemoradiotherapy plus sintilimab in pMMR/MSS rectal cancer patients with PD-L1 TPS ≥ 1% or CPS ≥ 1: an open-label, prospective, phase II study. npj Precis Oncol 9(1), 237 (2025). doi:10.1038/s41698-025-01018-0

28. Laengle J, Kuehrer I, Kulu A, et al. Dual Immune Checkpoint Inhibition Plus Neoadjuvant Chemoradiotherapy in Rectal Cancer: A Randomized Clinical Trial. JAMA Netw Open 8(8), e2527769 (2025). doi:10.1001/jamanetworkopen.2025.27769

29. Corgnac S, Malenica I, Mezquita L, et al. CD103⁺CD8⁺ TRM Cells Accumulate in Tumors of Anti-PD-1-Responder Lung Cancer Patients and Are Tumor-Reactive Lymphocytes Enriched with Tc17. Cell Rep Med 1(7), 100127 (2020). doi:10.1016/j.xcrm.2020.100127

30. Banchereau R, Chitre AS, Scherl A, et al. Intratumoral CD103⁺CD8⁺ T Cells Predict Response to PD-L1 Blockade. J Immunother Cancer 9(4), e002231 (2021). doi:10.1136/jitc-2020-002231

31. Virassamy B, Caramia F, Savas P, et al. Intratumoral CD8⁺ T Cells with a Tissue-Resident Memory Phenotype Mediate Local Immunity and Immune Checkpoint Responses in Breast Cancer. Cancer Cell 41(3), 585–601.e8 (2023). doi:10.1016/j.ccell.2023.01.004

