## Supplementary Materials for "Deep Learning Spatial Profiling of CD103⁺CD8⁺ T Cells and Survival in Rectal Cancer After Neoadjuvant Chemoradiotherapy"

#### Supplementary Tables

##### Supplementary Table S1. Clinicopathological characteristics stratified by stromal $CD103^{+}CD8^{+}$ T-cell density

| Characteristics | Median (range) or n | | |
|---|---|---|---|
| | Low (n =15) | High (n =25) | p value |
| Age (years) | 67 (56-80) | 69 (39-88) | 0.911 |
| Sex (Male/ Female) | 13/ 2 | 16/ 9 | 0.120 |
| Operative time (min) | 482 (319-952) | 444 (256-1138) | 0.570 |
| Blood loss (ml) | 330 (0-5345) | 135 (0-2368) | 0.147 |
| CEA after NACRT (≥ 5/ < 5 / unknown) (ng/ml) | 3/ 11 | 8/ 17 | 0.482 |
| CA19-9 after NACRT (≥ 37/ < 37/ unknown) (U/ml) | 4/ 10 | 7/ 18 | 0.970 |
| Histological type (Differentiated/ Undifferentiated) | 14/ 1 | 22/ 3 | 0.586 |
| ypStage (≥ III/ < III) | 8/ 7 | 4/ 21 | 0.013 |
| Pathological tumor response grade (Grade ≥ 2/ < 2) | 7/ 6 | 8/ 17 | 0.191 |
| Adjuvant chemotherapy (+/ -) | 6/ 9 | 12/ 13 | 0.622 |

Supplementary Table S2. Clinicopathological characteristics of the study cohort.

| Characteristics | Median (range) or n |
|---|---|
| Age (years) | 67.5 (39-88) |
| Sex (Male/ Female) | 29/ 11 |
| Operative time (min) | 477.5 (256-1138) |
| Blood loss (ml) | 224 (0-5345) |
| CEA after NACRT (≥ 5/ < 5 / unknown) (ng/ml) | 11/ 28/ 1 |
| CA19-9 after NACRT (≥ 37/ < 37/ unknown) (U/ml) | 11/ 28/ 1 |
| Histological type (Differentiated/ Undifferentiated) | 36/ 4 |
| ypStage (≥ III/ < III) | 12/ 28 |
| Pathological tumor response grade (Grade ≥ 2/ < 2) | 15/ 23 |
| Lateral pelvic lymph node dissection (+/ -) | 23/ 17 |
| Adjuvant chemotherapy (+/ -) | 18/ 22 |

Supplementary Table S3. Univariate and multivariate Cox proportional hazard regression models for relapse-free survival (RFS) and overall survival (OS) based on manual pathologist quantification

| | Univariate | Univariate | Multivariate-1 | Multivariate-1 | Multivariate-2 | Multivariate-2 | | | |
|---|---|---|---|---|---|---|---|---|---|
| Relapse-free survival | HR (95% CI) | p value | HR (95% CI) | p value | HR (95% CI) | p value | | | |
| Age (≥ 78/ < 78 y.o.) | 2.749 (0.786-9.619) | 0.114 | | | | | | | |
| Sex (M/ F) | 0.495 (0.142-1.730) | 0.271 | | | | | | | |
| Operative time (≥ 769/ < 769min) | 1.544 (0.502-4.753) | 0.449 | | | | | | | |
| Blood loss (≥ 315/ < 315mL) | 2.147 (0.791-5.829) | 0.134 | | | | | | | |
| CEA after NACRT (≥ 5/ < 5 ng/ml) | 0.624 (0.199-1.951) | 0.417 | | | | | | | |
| CA19-9 after NACRT (≥ 37/ < 37 U/ml) | 1.379 (0.476-3.992) | 0.554 | | | | | | | |
| ypStage (≥ III/ < III) | 2.737 (1.048-7.147) | 0.040 | 2.696 (1.014-7.168) | 0.047 | 2.137 (0.714-6.396) | 0.175 | | | |
| Pathological tumor response grade (Grade ≥ 2/ < 2) | 0.577 (0.2000-1.664) | 0.308 | | | | | | | |
| Adjuvant chemotherapy (+/ -) | 0.666 (0.253-1.757) | 0.412 | | | | | | | |
| Stromal CD8+ T-cell density by the pathologist (≥ 1022.5/ < 1022.5 cells/mm2) | 0.024 (0.000-1.515) | 0.078 | | | | | | | |
| Intratumoral CD8+ T-cell density by the pathologist (≥ 91/ < 91 cells/mm2) | 0.288 (0.108-0.773) | 0.013 | 0.290 (0.107-0.791) | 0.016 | | | | | |
| Stromal CD103+CD8+ T-cell density by the pathologist (≥ 8.5/ < 8.5 cells/mm2) | 0.230 (0.063-0.834) | 0.025 | | | 0.393 (0.092-1.692) | 0.210 | | | |
| Intratumoral CD103+CD8+ T-cell density by the pathologist (≥ 21.5/ < 21.5 cells/mm2) | 0.430 (0.162-1.139) | 0.089 | | | | | | | |
| | Univariate | Univariate | Multivariate-1 | Multivariate-1 | Multivariate-2 | Multivariate-2 | Multivariate-3 | Multivariate-3 | |
| Overall survival | HR (95% CI) | p value | HR (95% CI) | p value | HR (95% CI) | p value | HR (95% CI) | p value | p value |
| Age (≥ 75/ < 75 y.o.) | 5.049 (1.631-15.634) | 0.005 | 3.251 (0.987-10.714) | 0.053 | 4.766 (1.505-15.094) | 0.008 | 6.726 (2.006-22.544) | 0.002 | 0.002 |
| Sex (M/ F) | 0.199 (0.026-1.530) | 0.121 | | | | | | | |
| Operative time (≥ 400/ <400 min) | 1.298 (0.357-4.720) | 0.692 | | | | | | | |
| Blood loss (≥ 143/ <143 mL) | 1.853 (0.570-6.020) | 0.305 | | | | | | | |
| CEA after NACRT (≥ 5/ < 5 ng/ml) | 0.406 (0.089-1.860) | 0.246 | | | | | | | |
| CA19-9 after NACRT (≥ 37/ < 37 U/ml) | 2.205 (0.691-7.043) | 0.182 | | | | | | | |
| ypStage (≥ III/ < III) | 2.292 (0.768-6.838) | 0.137 | | | | | | | |
| Pathological tumor response grade (Grade ≥ 2/ < 2) | 0.304 (0.066-1.408) | 0.128 | | | | | | | |
| Adjuvant chemotherapy (+/ -) | 0.925 (0.210-2.756) | 0.888 | | | | | | | |
| Stromal CD8+ T-cell density by the pathologist (≥ 843.5/ < 843.5 cells/mm2) | 0.209 (0.046-0.949) | 0.043 | 0.302 (0.061-1.492) | 0.142 | | | | | |
| Intratumoral CD8+ T-cell density by the pathologist (≥ 352/ < 352 cells/mm2) | 0.037 (0.000-17.932) | 0.295 | | | | | | | |
| Stromal CD103+CD8+ T-cell density by the pathologist (≥ 12/ < 12 cells/mm2) | 0.270 (0.074-0.985) | 0.047 | | | 0.307 (0.082-1.157) | 0.081 | | | |
| Intratumoral CD103+CD8+ T-cell density by the pathologist (≥ 2.5/ < 2.5 cells/mm2) | 0.217 (0.058-0.807) | 0.023 | | | | | 0.143 (0.035-0.587) | 0.007 | 0.007 |

Bold values indicate statistical significance (p < 0.05).

Supplementary Table S4. Sensitivity analyses for relapse-free survival (RFS)

| Subgroup | n | HR (95% CI) for stromal CD103+CD8 TIL density | p value |
|---|---|---|---|
| All patients | 40 | 0.168 (0.057-0.494) | 0.001 |
| ypStage ≤ II | 28 | 0.461 (0.110-1.936) | 0.290 |
| ypStage ≥ III | 12 | 0.011 (0.000-3.752) | 0.130 |
| Adjuvant chemotherapy (+) | 18 | 0.210 (0.038-1.157) | 0.073 |
| Adjuvant chemotherapy (-) | 22 | 0.134 (0.032-0.555) | 0.006 |
| RFS ≥ 36 months | 22 | 0.000 (0.000-3.392E+14) | 0.689 |

Supplementary Figures

### Figure S1.

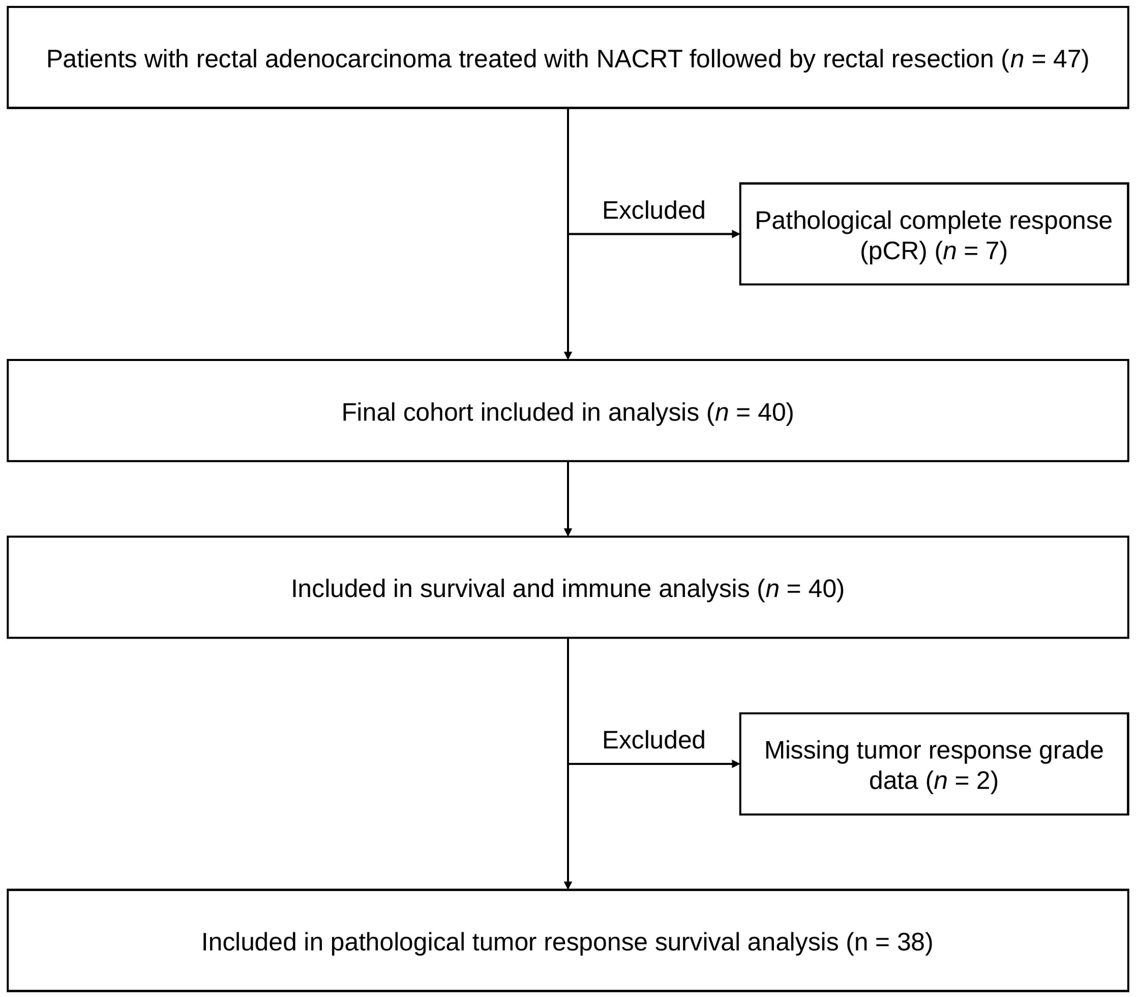

Figure S1. Study design and patient flow. Flow diagram showing patient selection. Of 47 patients with rectal adenocarcinoma treated with neoadjuvant chemoradiotherapy (NACRT) followed by radical resection, 7 who achieved pathological complete response were excluded, leaving 40 patients for analysis. Pathological tumor response was evaluable in 38 of these 40 patients.

### Figure S2.

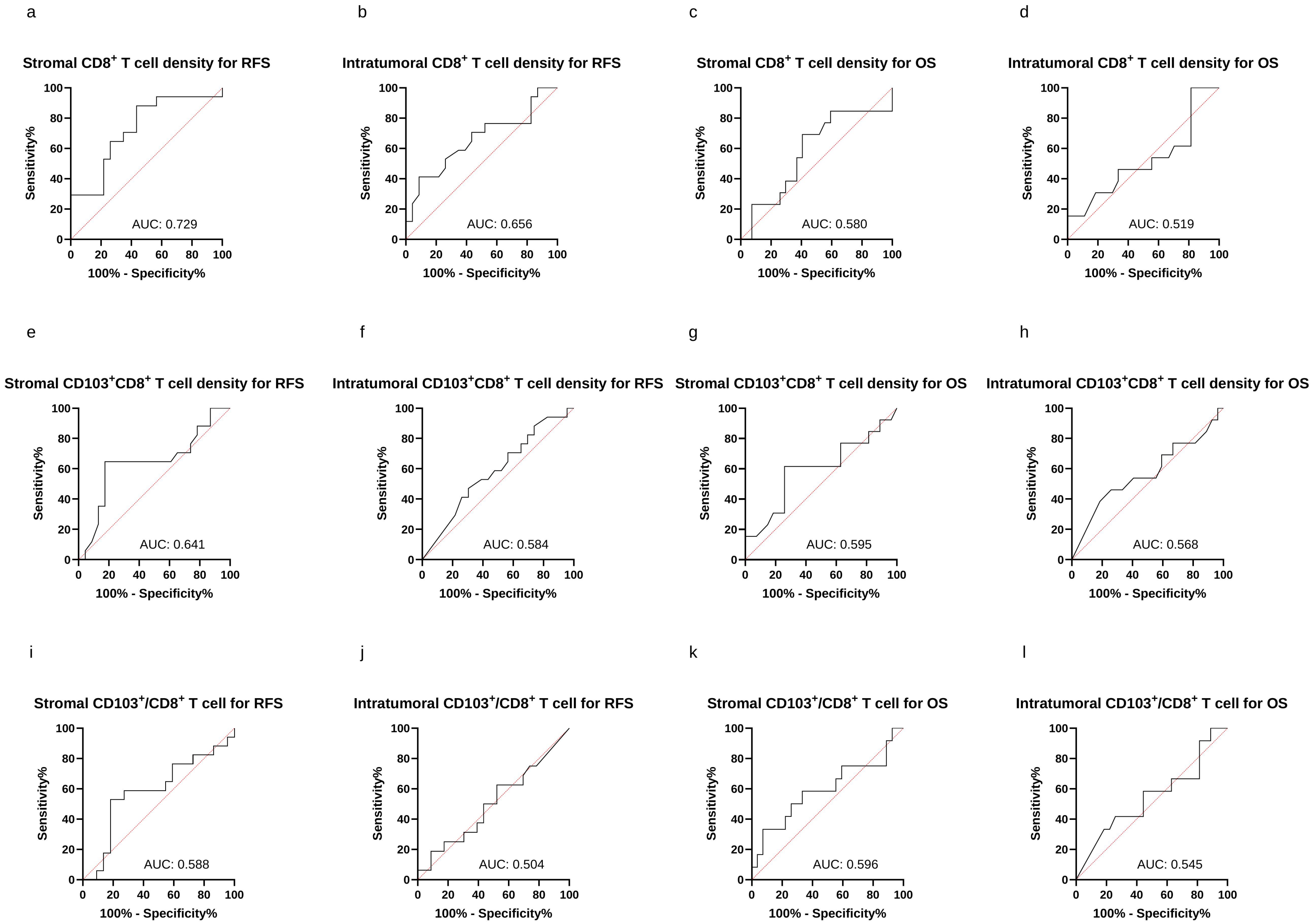

Figure S2. Receiver operating characteristic (ROC) curve analyses for determination of optimal cut-off values. ROC curve analyses were performed to determine the optimal cut-off values of stromal and intratumoral $CD8^+$ and $CD103^+CD8^+$ T-cell densities for predicting relapse-free survival (RFS) and overall survival (OS) in patients with rectal cancer after neoadjuvant chemoradiotherapy (NACRT). The areas under the curve (AUCs) for each parameter are shown. (a–d) ROC curves for $CD8^+$ T-cell density: (a) stromal $CD8^+$ for RFS, (b) intratumoral $CD8^+$ for RFS, (c) stromal $CD8^+$ for OS, (d) intratumoral $CD8^+$ for OS. (e–h) ROC curves for $CD103^+CD8^+$ T-cell density: (e) stromal $CD103^+CD8^+$ for RFS, (f) intratumoral $CD103^+CD8^+$ for RFS, (g) stromal $CD103^+CD8^+$ for OS, (h) intratumoral $CD103^+CD8^+$ for OS. (i–l) ROC curves for the proportion of $CD103^+$ cells among $CD8^+$ T cells: (i) stromal proportion for RFS, (j) intratumoral proportion for RFS, (k) stromal proportion for OS, (l) intratumoral proportion for OS.

### Figure S3.

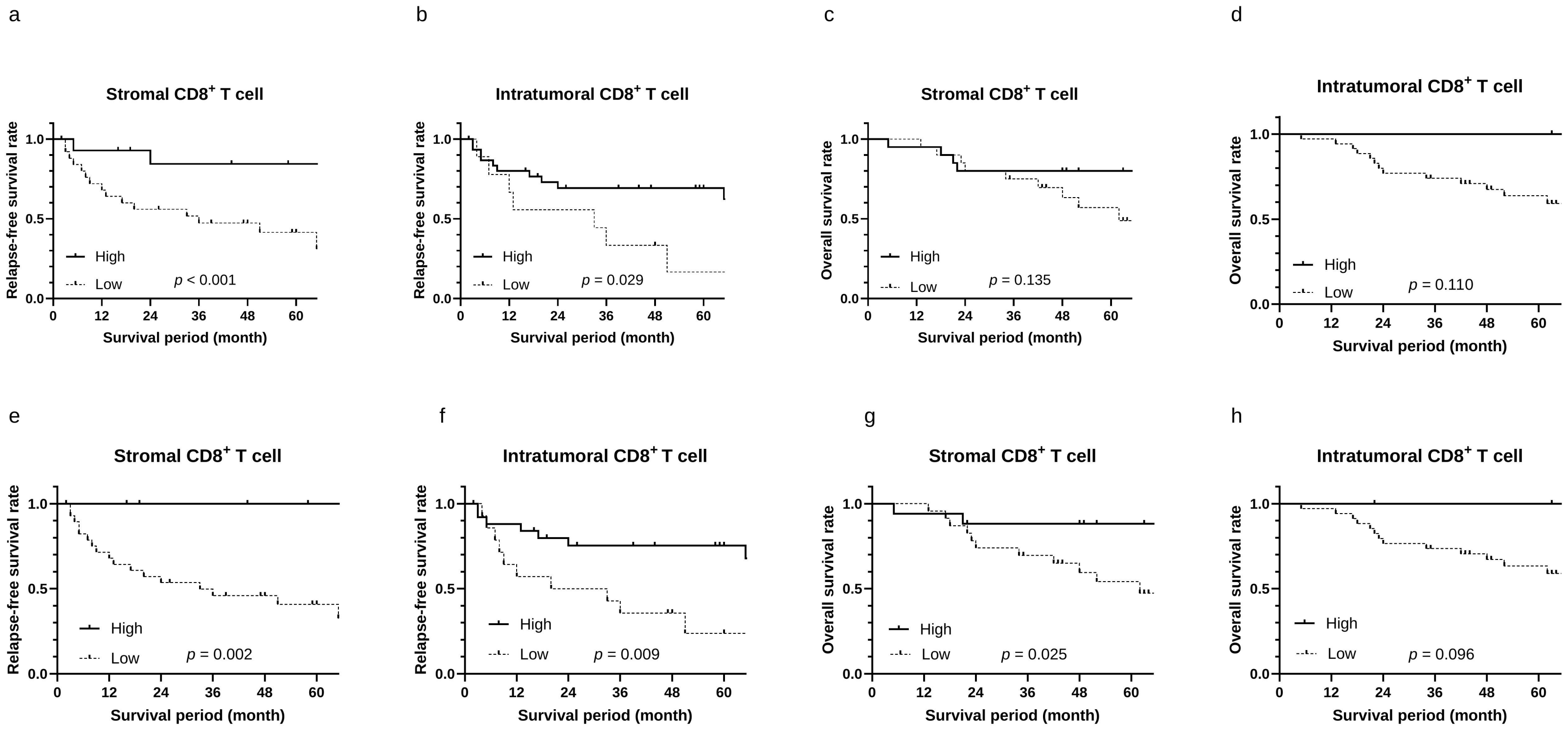

Figure S3. Prognostic significance of CD8⁺ T-cell density using Cu-Cyto versus manual pathologist quantification. Kaplan–Meier survival analyses were performed using CD8⁺ T-cell densities quantified by the deep learning-based Cu-Cyto platform (a–d) and manually by pathologists (e–h) in patients with rectal cancer after neoadjuvant chemoradiotherapy (NACRT). (a, e) Relapse-free survival (RFS) according to stromal CD8⁺ T-cell density, (b, f) RFS according to intratumoral CD8⁺ T-cell density, (c, g) overall survival (OS) according to stromal CD8⁺ T-cell density, and (d, h) OS according to intratumoral CD8⁺ T-cell density. High densities of both stromal and intratumoral CD8⁺ T cells were significantly associated with better RFS, showing high concordance between Cu-Cyto and manual quantification methods.

### Figure S4.

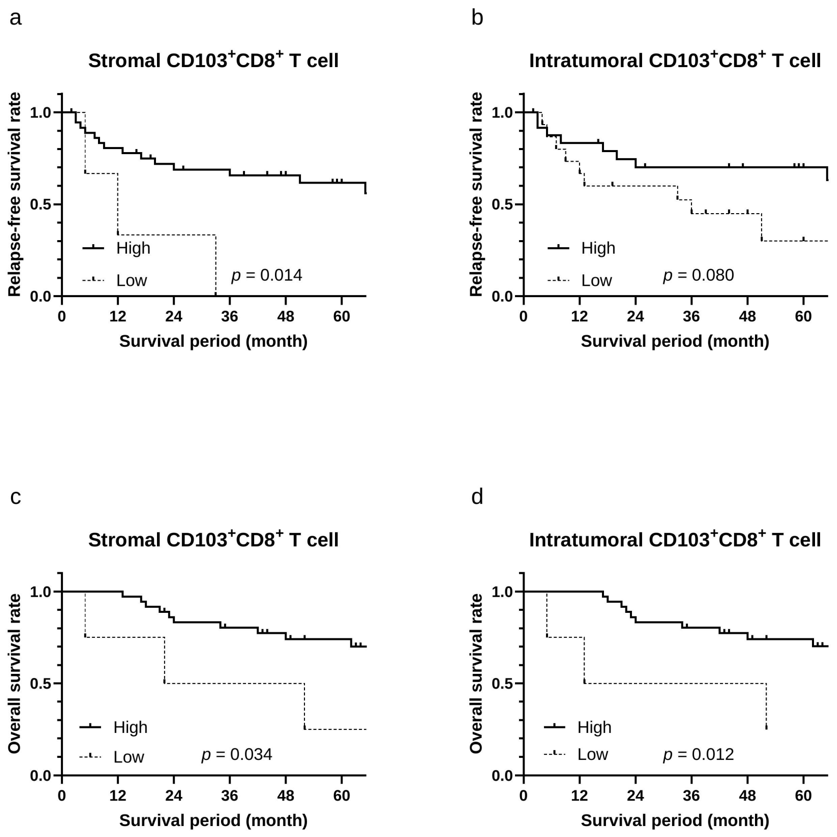

Figure S4. Kaplan–Meier survival analyses of $CD103^+CD8^+$ T cells based on manual quantification by pathologists. Kaplan–Meier survival analyses were performed using $CD103^+CD8^+$ T-cell densities manually quantified by pathologists in patients with rectal cancer after neoadjuvant chemoradiotherapy (NACRT). (a) Relapse-free survival (RFS) according to stromal $CD103^+CD8^+$ T-cell density, (b) RFS according to intratumoral $CD103^+CD8^+$ T-cell density, (c) overall survival (OS) according to stromal $CD103^+CD8^+$ T-cell density, and (d) OS according to intratumoral $CD103^+CD8^+$ T-cell density. The results were largely consistent with those obtained by Cu-Cyto–based quantification, supporting the robustness of stromal $CD103^+CD8^+$ T cells as prognostic biomarkers.

### Figure S5.

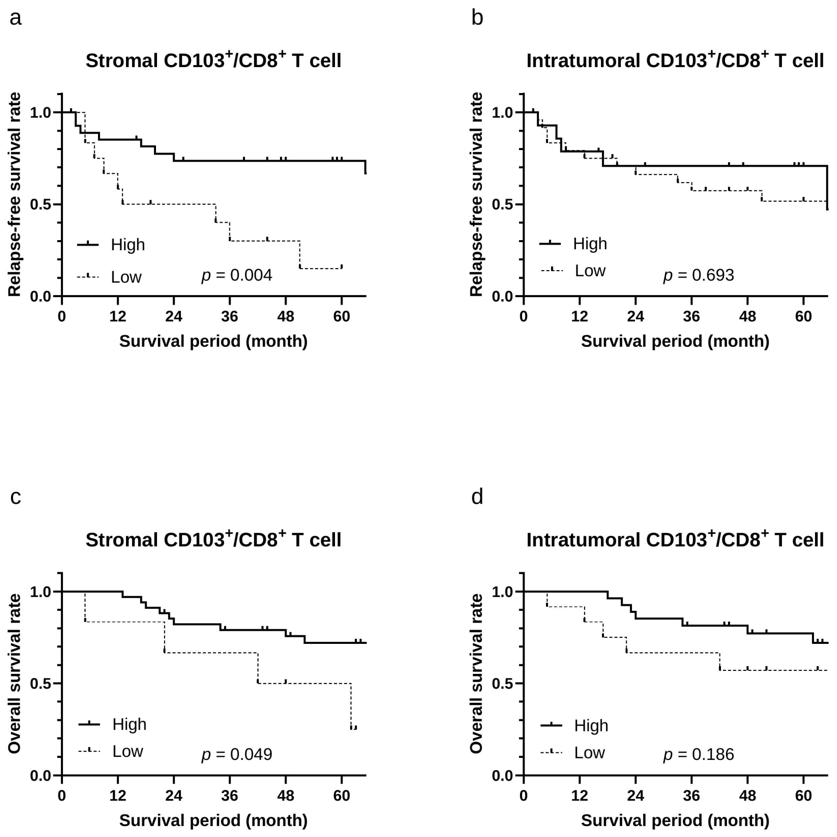

Figure S5. Kaplan–Meier survival analyses based on the proportion of CD103⁺ cells among CD8⁺ T cells. Kaplan–Meier survival analyses were performed using the proportion of CD103⁺CD8⁺ T cells among total CD8⁺ T cells, as quantified by Cu-Cyto, in patients with rectal cancer after neoadjuvant chemoradiotherapy (NACRT). (a, b) Relapse-free survival (RFS): (a) stromal proportion of CD103⁺ cells among CD8⁺ T cells and (b) intratumoral proportion of CD103⁺ cells among CD8⁺ T cells. (c, d) Overall survival (OS): (c) stromal proportion of CD103⁺ cells among CD8⁺ T cells and (d) intratumoral proportion of CD103⁺ cells among CD8⁺ T cells. In contrast to the absolute density of stromal CD103⁺CD8⁺ T cells, the proportion of CD103⁺ cells within CD8⁺ T cells did not show a significant association with either endpoint.
